# Cellular and humoral immunity towards parental SARS-CoV-2 and variants of concern after two doses of the NVX-CoV2373-vaccine in comparison to homologous BNT162b and mRNA1273 regimens

**DOI:** 10.1101/2022.08.02.22278342

**Authors:** Franziska Hielscher, Tina Schmidt, Verena Klemis, Alexander Wilhelm, Stefanie Marx, Amina Abu-Omar, Laura Ziegler, Candida Guckelmus, Rebecca Urschel, Urban Sester, Marek Widera, Martina Sester

## Abstract

The NVX-CoV2373-vaccine has recently been licensed, although data on vaccine-induced humoral and cellular immunity towards the parental strain and variants of concern (VOCs) in comparison to dual-dose mRNA-regimens are limited. In this observational study including 66 participants, we show that NVX-CoV2373-induced IgG-levels were lower than after vaccination with BNT162b2 or mRNA-1273 (n=22 each, p=0.006). Regardless of the vaccine and despite different IgG-levels, neutralizing activity towards VOCs was highest for Delta, followed by BA.2 and BA.1. Interestingly, spike-specific CD8 T-cell levels after NVX-CoV2373-vaccination were significantly lower and were detectable in 3/22 (14%) individuals only. In contrast, spike-specific CD4 T-cells were induced in 18/22 (82%) individuals. However, CD4 T-cell levels were lower (p<0.001), had lower CTLA-4 expression (p<0.0001) and comprised less multifunctional cells co-expressing IFNγ, TNFαα and IL-2 (p=0.0007) as compared to mRNA-vaccinated individuals. Unlike neutralizing antibodies, NVX-CoV2373-induced CD4 T cells cross-reacted to all tested VOCs from Alpha to Omicron, which may hold promise to protect from severe disease.

## Introduction

Among the COVID-19 vaccines, the NVX-CoV2373 vaccine was the first protein-based vaccine that was licensed as a homologous dual dose regimen in the European Union and in the United States [1-3]. NVX-CoV2373 is a prefusion-stabilized recombinant spike protein subunit vaccine with a saponin-based matrix M adjuvant that conferred a 89.7-90.4% protection towards parental SARS-CoV-2 infection, with a good safety profile and high efficacy against the B.1.1.7 variant [4, 5]. The efficacy exceeded those of the vector-based vaccines and was in a similar range as the two mRNA-based vaccines BNT162b2 and mRNA-1273, which were shown to be 95.0% and 94.1% efficacious, respectively, at preventing severe COVID-19 illness from the parental viral strains [6, 7].

The immunogenicity of the two mRNA vaccines has been extensively characterized in real world settings, and both vaccines induce strong antibody and T-cell responses [8, 9]. Likewise, the phase 1-2 trial of the NVX-CoV2373 vaccine or a recent phase III trial has shown induction of high antibody titers [10, 11], and first data on neutralizing activity towards SARS-CoV-2 variants of concern (VOCs) from participants of the pivotal trial 3-4 months after the second dose became available recently [12-14]. However, real world data on the immunogenicity of the NVX-CoV2373 vaccine during the induction phase, especially regarding its ability to induce CD4 and CD8 T cells and in comparison to other licensed vaccines are currently lacking. Given the fact that the vaccine was licensed during the surge of the delta and the omicron wave with its subvariants that are known to escape neutralizing antibody activity, knowledge of cellular immunity and its reactivity towards VOCs is of particular importance to inform on the ability to confer protection from severe disease.

We therefore carried out an observational study on the immunogenicity and reactogenicity of the NVX-CoV2373 vaccine in immunocompetent individuals who were vaccinated when the vaccine became approved and recommended in Germany [2]. Apart from vaccine-related adverse events, we characterized the induction of spike-specific IgG as well as CD4 and CD8 T cells including humoral and cellular reactivity towards the parental SARS-CoV-2 and variants of concern. Moreover, immune-responses after two doses of the vaccine were compared with respective data from matched cohorts of mRNA vaccinated individuals.

## Results

### Study population

This observational study included 22 healthy immunocompetent individuals, who received the NVX-CoV2373-vaccine as a standard two-dose regimen as per German recommendations [2]. Individuals after dual vaccination with BNT162b2 (n=22) and mRNA-1273 (n=22) matched for age and gender served as controls and were derived in part from convenience cohorts as described before [8, 9] (table 1). All individuals had no known history of SARS-CoV-2 infection and were negative for nucleocapsid-specific IgG. The mean time interval between the two vaccinations was 21.5±1.5 days for NVX-CoV2373, 31.2±9.5 days for BNT162b2, and 40.2±4.0 days for mRNA-1273, in accordance with German recommendations [2]. Blood sampling was carried out at a median of 15 (IQR 2) days after the second vaccination. Except for granulocytes, which were lower in individuals after BNT162b2 vaccination, the main leukocyte subpopulations did not differ between the groups (table 1). This also held true for numbers of monocytes, lymphocytes and lymphocyte subpopulations such as B cells, CD4 and CD8 T cells. Among B cells, plasmablast numbers, identified as CD38 positive cells among IgD^-^CD27^+^ CD19 positive switched-memory B cells were also similar in all groups (table 1).

**Table 1:**
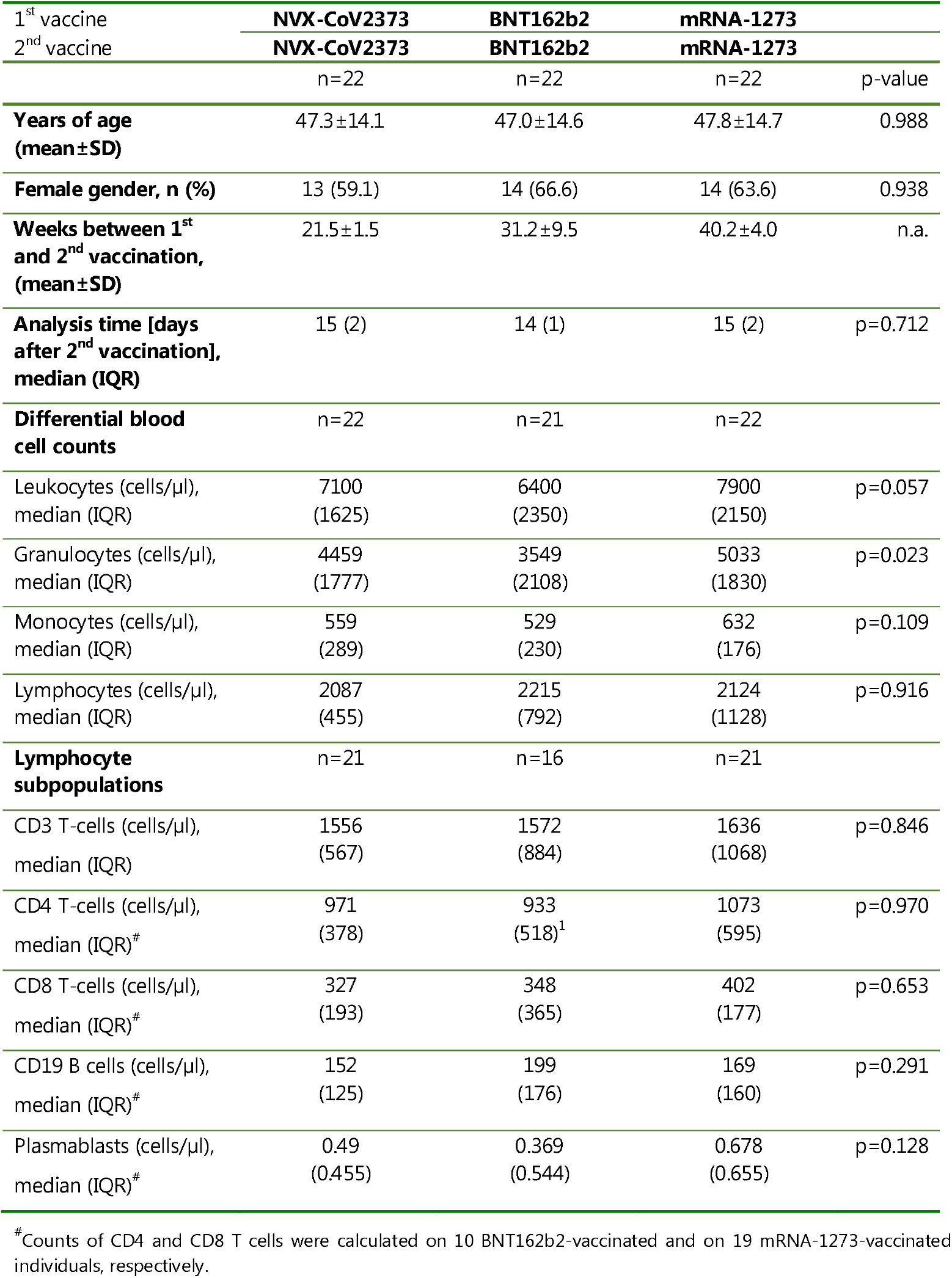
Demographic and basic characteristics of the study population.

### Evolution of antibodies and T cells after NVX-CoV2373 vaccination in individuals with and without prior infection

To study the evolution of NVX-CoV2373-induced anti-spike IgG, neutralizing antibodies and CD4 and CD8 T cells upon first and second vaccination, five individuals were studied before vaccination, as well as 14 days after the first and the second dose. In addition, four individuals with a history of one prior SARS-CoV-2 infection episode were analyzed before and 14 days after one single dose of the NVX-CoV2373-vaccine. Spike-specific IgG were quantified using ELISA and their neutralizing activity was determined using a surrogate assay. Spike-specific CD4 and CD8 T cells were quantified directly from whole blood samples after stimulation with overlapping peptides derived from the spike protein with diluent and Staphylococcus aureus Enterotoxin B (SEB) as negative and positive controls, respectively. Specifically stimulated CD4 and CD8 T cells were quantified using flow-cytometry based on the induction of the activation marker CD69 and the cytokines IFNγ, TNFαα or IL-2. Dotplots of a representative example of spike-specific CD4 and CD8 T-cell analyses of a 37-year-old female after the second vaccination with respective control stimulations are shown in figure 1A. As shown in figure 1B, no specific immunity was detectable prior to vaccination in infection-naïve individuals, and the first vaccine dose only led to detectable specific antibodies, neutralizing activity and T cells in a subset of individuals. In contrast, anti-spike IgG were induced in all individuals after the second dose with a significant increase in IgG-levels and neutralizing activity. As shown for CD69+IFNγ+ T cells, the second vaccine-dose induced spike-specific CD4 T cells to a variable extent, whereas CD8 T cells were largely absent (figure 1B). In all individuals with prior infection, one dose of the vaccine readily induced IgG, strong neutralizing activity and CD4 T cells, whereas spike-specific CD8 T cells remained below detection limit (figure 1C). Polyclonally stimulated CD4 and CD8 T-cell levels remained constant throughout the observation period (figure 1B and C, bottom panels).

**Figure 1:**
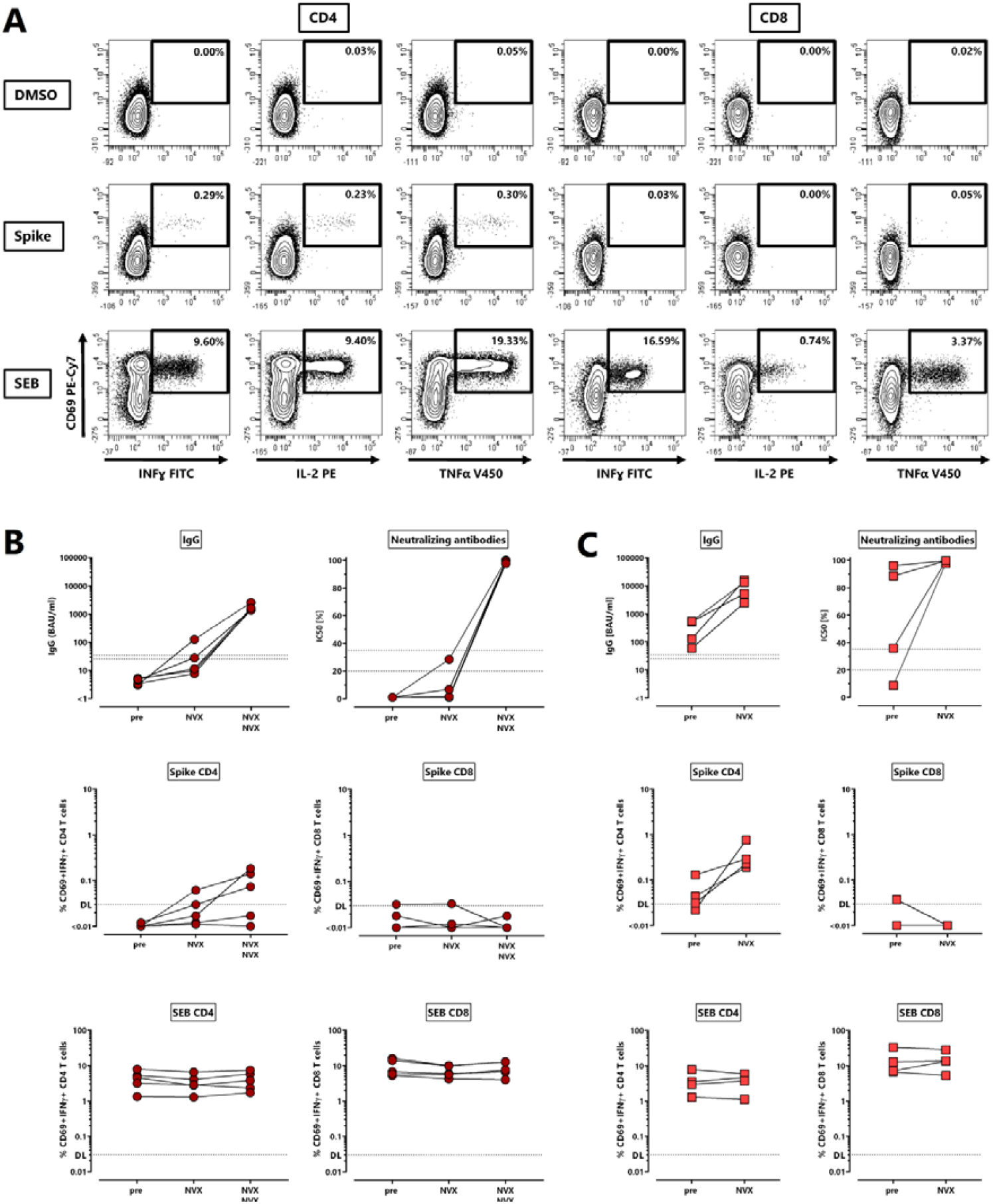
Evolution of antibodies and T cells against the SARS-CoV-2 spike protein after NVX-CoV2373-vaccination. **(A)** Representative contour plots of CD4 and CD8 T cells after stimulation of a whole blood sample from a 47-years old female 14 days after the second dose of the NVX-CoV2373-vaccine (NVX). Percentages of activated (CD69+) cells producing IFNγ, TNFαα or IL-2 among CD4 or CD8 T cells are shown after stimulation with DMSO (negative control), overlapping peptides of SARS-CoV-2 spike protein or SEB (positive control). Spike-specific IgG, neutralizing activity, spike-specific CD4 and CD8 T cells, and SEB-reactive CD4 and CD8 T cells were determined **(B)** in non-infected individuals (n=5) prior to vaccination as well as after the first and the second vaccination, or **(C)** in individuals with history of one prior infection (n=4) before and after a single dose of NVX-CoV2373-vaccine. Specific T cells in panels B and C show CD4 or CD8 T cells co-expressing CD69 and IFNγ with respective reactivity after control stimulation subtracted. Bars represent medians with interquartile ranges. Dotted lines represent detection limits for antibodies indicating negative, intermediate and positive levels or levels of inhibition, respectively as per manufacturer’s instructions, and detection limits for specific T cells. IFN, interferon; IL, interleukin; SEB, *Staphylococcus aureus* enterotoxin B; TNFα, tumor necrosis factor.

### Lower levels of spike-specific antibodies and T cells after vaccination with NVX-CoV2373 compared to dual-dose mRNA-vaccines

In the whole cohort of 22 individuals, the NVX-CoV2373-vaccine induced spike-specific IgG in all individuals (table 2), although median IgG levels after NVX-CoV2373-vaccination were significantly lower (2633 (IQR 3566) BAU/ml) as compared to individuals after homologous BNT162b2 or mRNA-1273 vaccination ((4870 (IQR 3414) BAU/ml) and (4932 (IQR 6686) BAU/ml), respectively, figure 2A, p=0.006). In contrast, neutralizing antibody activity against the parental spike protein which was determined by a surrogate assay was similarly high in all vaccine groups (figure 2A). To further characterize neutralizing activity of vaccine-induced antibodies towards the authentic SARS-CoV-2 VOCs Delta and Omicron BA.1 and BA.2, a micro-neutralisation assay was performed. As shown in table 2, the percentage of individuals with neutralizing activity towards Delta was highest (17/22 after NVX-CoV2373-vaccination, and 21/22 after BNT162B2- and mRNA-1273 vaccination), followed by BA.2 (25-50% of individuals), whereas activity towards BA.1 was largely absent. However, despite significant differences in anti-spike IgG-levels, median IC_50_-titers did not differ between the vaccine groups (figure 2B). Despite the small sample size of four individuals with prior infection only, it was remarkable that one dose of NVX-CoV2373-induced neutralizing activity towards all VOCs with in part markedly higher IC_50_-levels (figure 3C).

**Table 2:** Qualitative results of antibody and T-cell induction after the second vaccination.

| 1° vaccine | NVX-CoV2373 | BNT162b2 | mRNA-1273 |  |
| --- | --- | --- | --- | --- |
| 2° vaccine | NVX-CoV2373 | BNT162b2 | mRNA-1273 |  |
| n (% positive) | n=22 | n=22 | n=22 | p-value |
| **IgG** ($\geq 35.2$ BAU/ml) | 22 (100%) | 22 (100%) | 22 (100%) | p=1.00 |
| **Neutralizing activity** |  |  |  |  |
| **Parental** (IH$\geq 35\%$)<sup>1</sup> | 22 (100%) | 22 (100%) | 22 (100%) | p=1.00 |
| **Delta** (>0%)<sup>2</sup> | 17 (77.3%) | 21 (95.5%) | 21 (95.5%) | p=0.07 |
| **BA.1** (>0%)<sup>2</sup> | 1 (4.5%) | 0 (0%) | 4 (18.2%) | p=0.06 |
| **BA.2** (>0%)<sup>2,3</sup> | 8 (36.4%) | 4 (25%)<sup>1</sup> | 11 (50.0%) | p=0.29 |
| **CD4 T cells** ($\geq 0.03\%$) | 18 (81.8%) | 22 (100%) | 22 (100%) | **p=0.01** |
| **CD8 T cells** ($\geq 0.03\%$) | 3 (13.6%) | 14 (63.6%) | 17 (77.3%) | **p<0.0001** |

<sup>1</sup>determined by a surrogate assay; <sup>2</sup>determined by a live viral assay; <sup>3</sup>BA.2 testing only available for n=16 individuals. Shown is the number (percentage) of individuals with specific immune responses above the respective detection limit.

**Figure 2:**
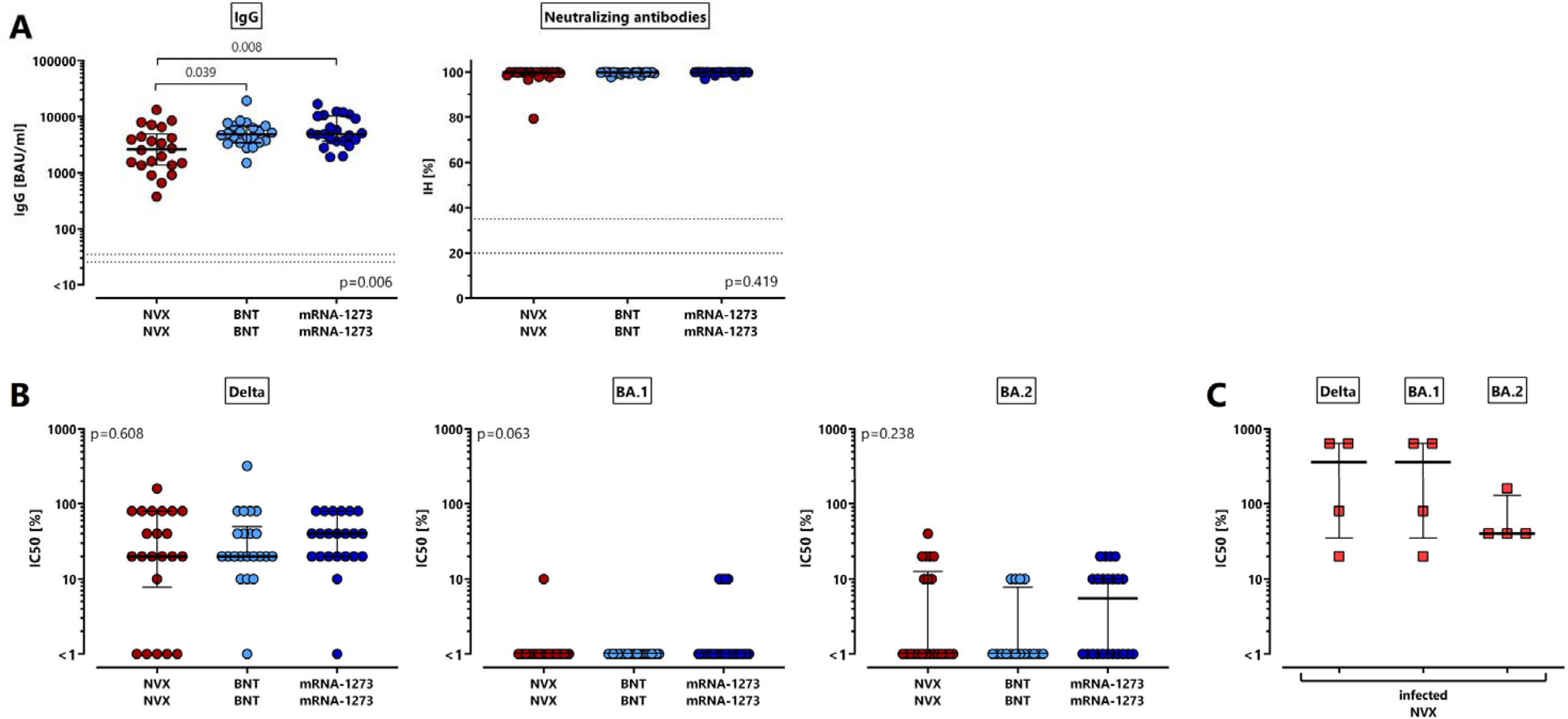
Lower levels of spike-specific IgG after NVX-COV2373-vaccination with similar neutralizing activity as mRNA-vaccinated individuals. Spike-specific antibodies were characterized among 66 individuals 13-18 days after the second vaccination with NVX-CoV2373 (NVX), BNT162b2 (BNT) or mRNA-1273 (n=22 each). **(A)** ELISA and a surrogate neutralization assay were performed to quantify levels of spike-specific IgG and neutralizing antibodies towards parental SARS-CoV-2 (expressed as percentage of inhibition (IH)). **(B)** Antibody-mediated neutralisation of authentic SARS-CoV-2 variants of concern Delta and Omicron BA.1 and BA.2 variants of concern was tested among the three vaccine groups and **(C)** in the four individuals with history of infection after the first NVX-COV2373-vaccine dose. Bars represent medians with interquartile ranges (expressed as IC_50_). Differences between the groups were calculated using two-sided Kruskal-Wallis test with Dunn’s multiple comparisons post-test. Dotted lines represent detection limits indicating negative, intermediate and positive IgG-levels or levels of inhibition, respectively as per manufacturer’s instructions..

**Figure 3:**
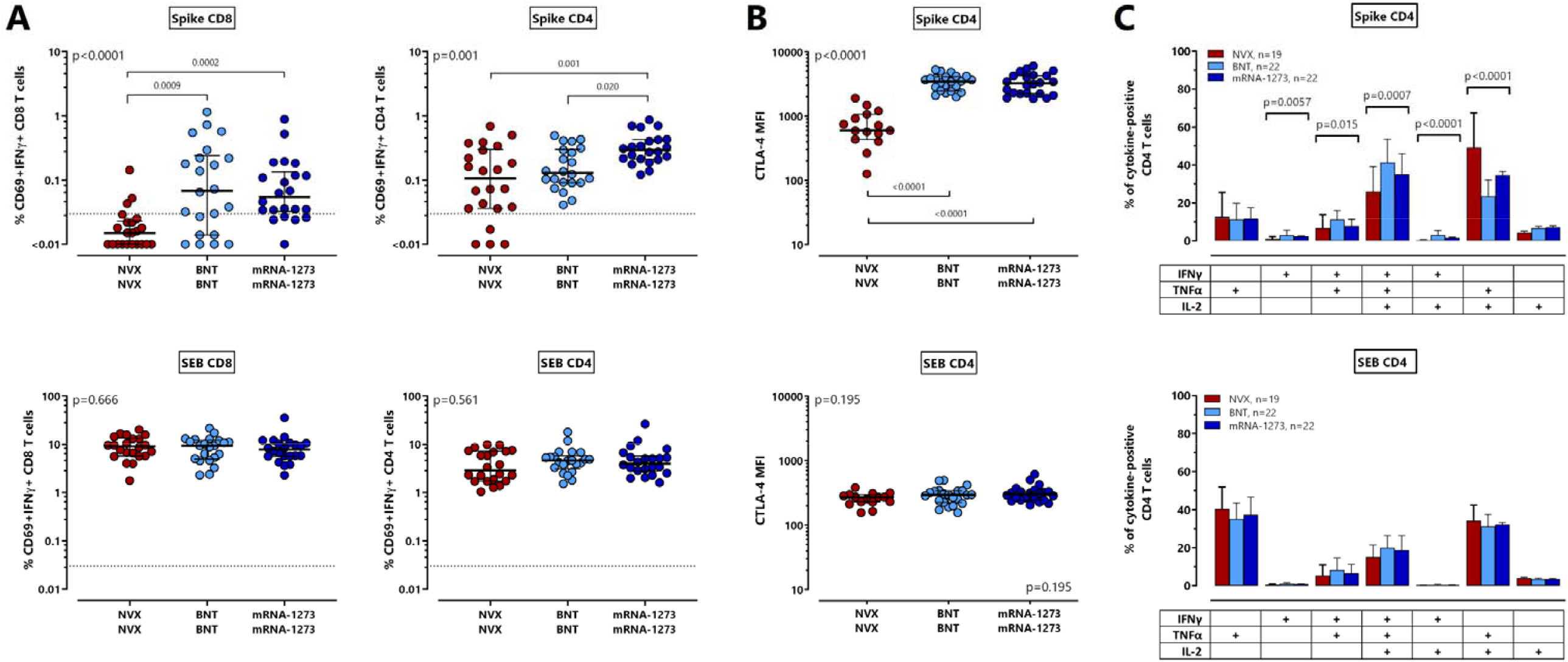
Lower levels of spike-specific T cells, lower CTLA-4 expression and lower percentages of polyfunctional CD4 T cells after NVX-COV2373-vaccination. Spike-specific CD4 and CD8 T cells were quantified and characterized by intracellular cytokine staining after antigen-specific stimulation of whole blood samples of 66 individuals 13-18 days after the second vaccination with NVX-CoV2373 (NVX), BNT162b2 (BNT) or mRNA-1273 (n=22 each). **(A)** Spike-specific and SEB-reactive CD4 and CD8 T cells were determined based on co-expression of CD69 and IFNγ with respective reactivity after control stimulation subtracted. Bars represent medians with interquartile ranges. Dotted lines indicate detection limits for specific T cells. **(B)** CTLA-4 expression of spike-specific and SEB-reactive CD4 T cells was determined in all samples with at least 20 CD69+ IFNγ-positive CD4 T cells (n=15 for NVX CoV2373, n=22 for mRNA-vaccinated individuals). Bars in panels A and B represent medians with interquartile ranges. Differences between the groups were calculated using two-sided Kruskal-Wallis test with Dunn’s multiple comparisons post-test. **(C)** After spike-specific or polyclonal stimulation, cytokine expressing CD4 T cells were subclassified into 7 subpopulations according to single or combined expression of IFNγ, TNFαα and IL-2. Blood samples from all individuals were analyzed. To ensure robust statistics, only samples with at least 30 cytokine-expressing CD4 T cells after normalization to the negative control stimulation were considered (with sample size in each vaccine group indicated in the figures). Bars represent means and standard deviations, and ordinary one-way ANOVA tests were performed. Analyses in panels B and C were restricted to CD4 T cells due to the lack of spike-specific CD8 T cells in most NVX-COV2373-vaccinated individuals. CTLA-4, cytotoxic T-lymphocyte associated protein 4; IFN, interferon; IL, interleukin; MFI, median fluorescence intensity; SEB, Staphylococcus aureus enterotoxin B; TNFα, tumor necrosis factor.

As with anti-spike IgG-levels, median percentages of spike-specific CD69+IFNγ+ CD4 and CD8 T cells were significantly lower after NVX-CoV2373-vaccination (figure 3A). The difference was most pronounced for vaccine-induced CD8 T-cell levels (figure 3A, p<0.0001), where only 3/22 (13.6%) individuals showed detectable CD8 T cells towards spike after NVX-CoV2373-vaccination. This contrasts with mRNA-regimens which induced specific CD8 T cells in most individuals (table 2, p<0.0001). Spike-specific CD4 T cells were induced in 18/22 (81.8%) NVX-CoV2373-vaccinated individuals, although their median levels were significantly lower (0.11% (IQR 0.27%)) as compared to individuals after vaccination with BNT162b2 or mRNA-1273 (0.13% (IQR 0.21%) and 0.29% (IQR 0.22%), respectively, p=0.001, figure 3A), where all had detectable CD4 T cells (table 2). Vaccine-induced CD4 T cells were further characterized for phenotypical and functional properties. In line with a less pronounced induction of vaccine-induced T-cell levels, spike-specific CD4 T cells after NVX-CoV2373-vaccination showed a significantly lower expression level of CTLA-4 as compared to both mRNA-groups, which may result from a lower extent of antigen encounter during the induction phase (figure 3B, p<0.0001). When spike-specific CD69 positive cells were tested for induction of other Th1 cytokines such as TNFαα or IL-2, results were largely similar as with IFNγ producing cells (supplementary figure S1). In addition, Boolean gating was used to analyze cytokine-expression profiles of spike-specific CD4 T cells producing IFNγ, TNFαα and IL-2 alone or in combination. As shown in figure 3C, specific CD4 T cells after NVX-CoV2373-vaccination comprised the lowest percentage of polyfunctional T cells simultaneously expressing IFNγ, TNFαα and IL-2, and a concomitant higher percentage of cells producing TNFαα and IL-2. All effects were spike-specific, as SEB-reactive CD4 or CD8 T-cell levels including their CTLA-4 expression and cytokine-expression profiles were similar in all groups (figure 3, bottom panels).

### NVX-CoV2373-induced T cells equally recognize parental SARS-CoV-2 and variants of concern

To analyze whether NVX-CoV2373-induced CD4 T cells were able to recognize antigens from variants of concern, we used UV-inactivated viruses including the parental strain (D614G, FFM7) and the VOCs Alpha, Beta, Delta, and Omicron BA.1 and BA.2 to stimulate vaccine-induced T cells *in vitro*. As shown for two representative individuals with detectable spike-specific CD4 T-cell immunity, all VOCs elicited a similar percentage of specific CD4 T cells as the parental strain (figure 4A). Moreover, the percentage of CD4 T cells after stimulation with overlapping peptides derived from wildtype spike showed a significant correlation with T-cell levels obtained after stimulation with the UV-inactivated parental strain (r=0.92, p<0.0001), which indicates that the viral preparations were suitable for stimulation. Data from all individuals show that the T-cell frequencies reacting towards the parental strain within one individual largely correspond to the respective T-cell levels after stimulation with the variants (figure 4B). As shown in the heatmap in figure 4C, all pair-wise comparisons of specifically stimulated CD4 T-cell levels showed a strong, highly significant correlation.

**Figure 4:**
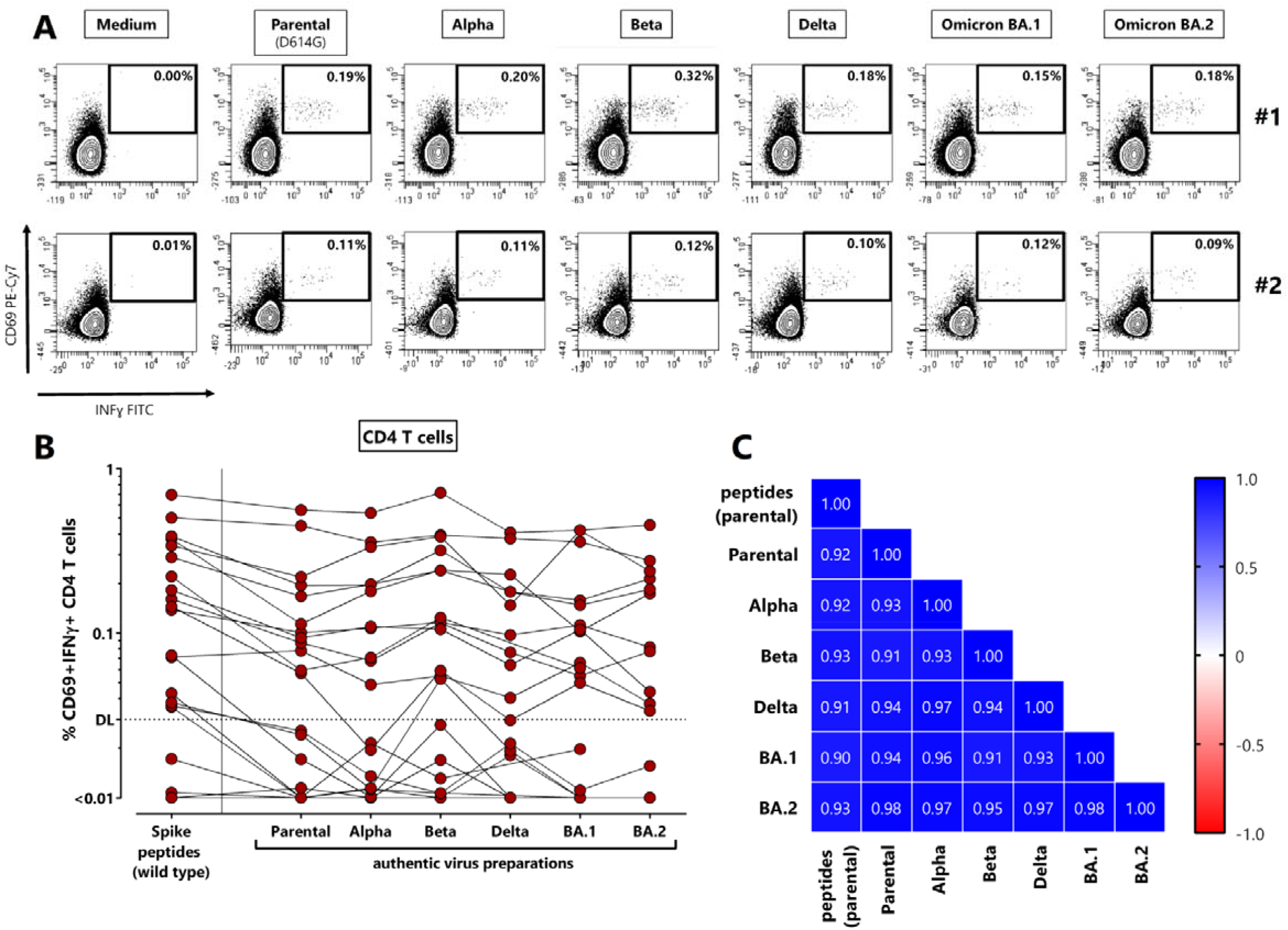
NVX-CoV2373-induced T cells equally recognize parental SARS-CoV-2 and variants of concern. Whole blood samples of 22 NVX-COV2373-vaccinated individuals 13-18 days after the second vaccination were stimulated with overlapping peptides of parental SARS-CoV-2 spike or UV-inactivated parental SARS-CoV-2 or variants of concern. **(A)** Dotplots of two individuals with detectable spike-specific CD4 T cells are shown (#1: 56 year old female, #2: 32 year old male). Percentages of activated (CD69+) cells producing IFNγ among CD4 T cells are shown after stimulation with medium (negative control), parental SARS-CoV-2 or variants Alpha, Beta, Delta, Omicron BA.1 and BA.2. **(B)** Specific CD4 T cells were quantified for all individuals based on co-expression of CD69 and IFNγ with respective reactivity after control stimulation subtracted. Indeterminate results of one person had to be excluded due to excessive background reactivity in the medium control; Delta and BA.2 stimulations were available from 19 individuals only. **(C)** Correlation matrix of specific T-cell levels determined after stimulation with spike-peptides and the different UV-inactivated virus preparations. Correlation coefficients were calculated according to two-tailed Spearman and displayed using a color code. IFN, Interferon;

### The NVX-CoV2373-vaccine was well tolerated with no striking differences in adverse events between the vaccine groups

Vaccine-related adverse events after the first and the second vaccination were self-reported using a questionnaire. The percentage of individuals reporting local or systemic adverse events after the first and the second vaccination was similar for all three vaccine groups (figure 5A). Regardless of the vaccine, the largest fraction of individuals reported of having been more affected by the second vaccination than by the first (figure 5B). Among local adverse events, pain at the injection site was more often reported than swelling (figure 5C) with no significant difference between the groups. Systemic adverse events including fever, headache, chills, gastrointestinal manifestations, and myalgia were similar for all vaccine groups. Arthralgia was significantly less frequently reported after the second BNT162B2-vaccination (p=0.005), whereas fatigue was least frequent after the second NVX-COV2373-vaccination (p=0.032). Overall, the need for antipyretic medication was similarly low in all groups. The notion that most adverse events were numerically least frequent after NVX-COV2373-vaccination indicates that this vaccine was at least equally well or better tolerated as the mRNA vaccines.

**Figure 5:**
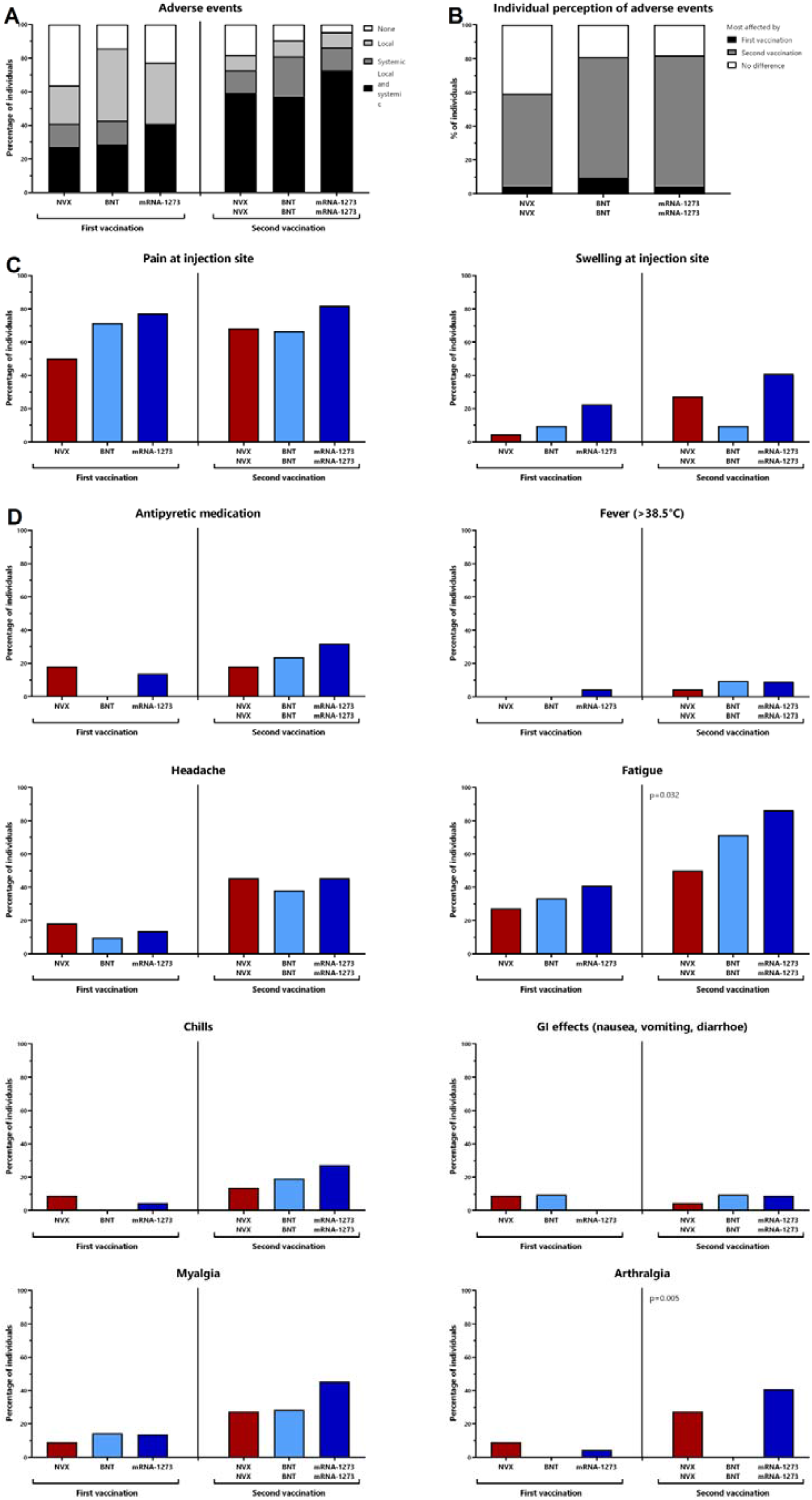
Reactogenicity after primary and secondary vaccination with NVX-CoV2373, BNT162b2or mRNA-1273. Self-reported reactogenicity within the first week after the first and the second vaccine dose was assessed using a standardized questionnaire. **(A)** The presence of local or systemic adverse events or both in general, **(B)** individual perception of which of the two vaccinations affected more, **(C)** local adverse events, or (D) systemic adverse events are shown. Statistical analysis was performed using X^2^ test. BNT, BNT162b2 vaccine; NVX, NVX-CoV2373 vaccine.

## Discussion

NVX-CoV2373 was recently licensed in Europe and in the United States as a dual dose regimen [1, 3], but immunogenicity data regarding cellular immunity and reactivity towards variants of concern are scarce. Based on a convenience cohort in a real-world setting we show that the vaccine was well tolerated and all individuals mounted a strong anti-spike IgG-response after two doses. Despite lower IgG-levels as compared to two doses of the mRNA-vaccines BNT152b2 or mRNA-1273, neutralizing activity towards the SARS-CoV-2 VOCs Delta and Omicron BA.1 and BA.2 VOCs was similar in all three vaccine groups. Of note, in individuals with prior history of infection, one dose of NVX-CoV2373 was sufficient to induce similar anti-spike IgG-levels with high neutralizing activity towards all VOCs as two doses in infection naïve individuals. In contrast to mRNA vaccines, the protein-based NVX-COV2373-vaccine poorly induced specific CD8 T cells, whereas spike-specific CD4 T cells were mounted in most individuals. Unlike poor neutralizing ability of antibodies towards VOCs, the magnitude of the CD4 T-cell response towards the VOCs was similar to the parental strain. Given that licensing of NVX-CoV2373 coincided with the circulation of VOCs such as Delta and Omicron, knowledge on the neutralizing activity of antibodies and of T-cell reactivity towards VOCs is of particular importance in providing information on the ability to confer protection against infection and severe disease.

In line with results from our study, recent data including seven different vaccine regimens have shown a severe dampening of neutralizing antibody activity towards VOCs as compared to the parental virus in all tested vaccine groups [12-14]. These studies also included ten individuals after NVX-COV2373-vaccination who participated in the randomized pivotal trial. However, the mean intervals between the last vaccination and analysis varied from 13 days in the two mRNA vaccine groups to up to 82 days in individuals after NVX-COV2373-vaccination [14]. Given the known dynamics in antibody titers over time, samples in some groups including NVX-CoV2373 were not captured at peak titers. Thus, although the neutralizing activity towards VOCs was consistently lower within one vaccine group, comparisons between vaccine groups were difficult. In our study, sample size was larger and analysis time was standardized in all tested groups, which allows for better head-to-head comparisons of antibody titers and neutralizing function between vaccine regimens.

Up to now, evidence for induction of NVX-CoV2373-induced memory T cells exist from 12 individuals tested 3-4 months after the second vaccination [12]. However, no T-cell data were available on NVX-CoV2373-induced T cells during the induction phase in comparison with other vaccine regimens. We now show that spike-specific CD4 T cells were readily induced in most individuals, albeit at lower levels as after mRNA-1273-vaccination. Lower CD4 T-cell levels were associated with a significantly lower expression of CTLA-4 and a lower percentage of multifunctional cells. We have previously shown that the phenotypical and functional profile of antigen-specific T cells correlated with antigen-load during active infections [15, 16] or vaccination [8, 9, 17] to counteract excessive T-cell proliferation or immunopathology. Therefore, the phenotypical and functional differences may indicate a lower extent of antigen encounter during the induction phase in NVX-CoV2373-vaccinated individuals. However, the most striking difference between NVX-CoV2373- and mRNA-vaccinated individuals was the poor induction of spike-specific CD8 T cells, which may result from the fact that proteins taken up by antigen-presenting cells during the induction phase of the immune response are less efficiently processed for presentation in MHC class I molecules. This contrasts with mRNA vaccines that rely on translation of the spike protein within the host cells [18]. This enables presentation of spike-derived peptides in both MHC class I and class II molecules which may explain the differences in the ability to induce CD4 and CD8 T cells by the different vaccine platforms.

While neutralizing antibodies have been associated with protection from infection, vaccine-induced T cells seem to play a major role in protecting from severe disease. Unlike neutralizing antibody activity, NVX-CoV2373-induced CD4 T cells analyzed two weeks after the second vaccination show considerable cross-reactivity towards all tested VOCs. This is consistent with findings from individuals after BNT162b2 or Ad26.CoV2.S vaccination during the induction phase [19] or memory T cells from eight NVX-CoV2373-vaccinated individuals tested 3-4 months after vaccination [13]. Thus, despite the current surge in the incidence of Omicron infections due to impaired neutralizing activity, the induction of T cells with robust cross-reactivity towards VOCs across several vaccine regimens likely plays a major role in retaining protection from severe disease. In this regard, the Th1 polarized phenotype of NVX-CoV2373-induced CD4 T cells with the ability to produce IFNγ, TNFαα and IL-2 may be instrumental in cell-mediated elimination of infected cells [20, 21]. Future effectiveness studies should determine whether the differences in CD4 T-cell levels and functionality and the relative lack of vaccine-induced CD8 T cells may reveal differences between vaccine regimens regarding disease severity upon breakthrough infection. Moreover, it will be interesting to study whether heterologous boosting with mRNA vaccines will induce specific CD8 T cells as was shown for heterologous mRNA-boosting after vector-vaccination [8, 9, 22-25]. Regarding neutralizing activity, a third booster dose after NVX-CoV2373 may seem equally valuable to improve protection as has been shown for other primary vaccine regimens [26-28]. This is supported by promising data from a small study showing that neutralizing antibody titers towards VOCs increased in all seven NVX-CoV2373-vaccinated individuals who either received mRNA-1273, BNT162b2 or Ad26.COV.2 as a booster dose [29].

A strength of our study is the head-to-head analysis of immunogenicity and reactogenicity of the NVX-COV2373-regimen in a real world setting in comparison with the two mRNA-regimens BNT162b2 and mRNA-1273, which are most widely used in European countries and many parts of the world [30]. Our study is limited by convenience sampling in a non-randomized study design, where the study participants on the various vaccine regimens were not enrolled in the same time frame. However, as individuals with evidence for prior infection were excluded from the main analysis, our results are not confounded by differences in circulating viral strains over time. Moreover, participants were enrolled according to national recommendations, where some vaccine-specific differences in the intervals between the first and the second vaccination may have an influence on the level of specific immunity. However, recent evidence suggests that a short interval in the range of 3-6 weeks as in our study does not appear to have an impact on immunogenicity [31]. Finally, information on the neutralizing activity or T-cell reactivity on the BA.4 and BA.5 variant is not available. However, neutralizing activity towards BA.4 and BA.5 in individuals 3-4 months after NVX-CoV2373-vaccination has recently been shown to be markedly lower than for BA.1 or BA.2, which has been equally observed with other vaccine regimens [29].

Occurrence of serious adverse events such as thrombosis [32, 33] or myocarditis [34] after vector-or mRNA-based vaccination has resulted in vaccine hesitancy that may be overcome by the availability of traditional vaccine formulations based on recombinant proteins. No such adverse events were described for NVX-CoV2373 so far and the vaccine was well tolerated by all participants of our study. Together with the ability to induce a potent antibody and CD4 T-cell response and practical advantages such as vaccine stability during refrigeration, NVX-CoV2373 may have potential for widespread use as either primary series or as booster vaccine pending further regulatory approval.

## Methods

### Study design and subjects

Study participants for NVX-CoV2373 vaccination were enrolled prospectively prior to the start of a vaccination cycle or no longer than 13-18 days after the second vaccination as described before [9]. In addition, individuals after dual dose vaccination with BNT162b2 and mRNA-1273 matched for age and sex were chosen as control groups. Their data and frozen plasma samples were in part derived from previous observational studies [8, 9]. The time interval between the first and the second vaccination (3 weeks for NVX-CoV2373, 3-6 weeks for mRNA regimens) was based on recommendations [2, 35] and not determined by the study. Study participants completed a questionnaire for self-reporting of local and systemic adverse events occurring within the first week after the first and second vaccination, respectively. Blood samples were collected during an interval of 13-18 days after secondary vaccination, or prior to vaccination or after the first vaccination in subgroups before and after NVX-CoV2373-vaccination. The study was approved by the ethics committee of the Ärztekammer des Saarlandes (reference 76/20), and all individuals gave written informed consent.

### Quantification of lymphocyte populations and plasmablasts

T cells, B cells and plasmablasts were quantified from 100 μl heparinized whole blood as described before [9] using monoclonal antibodies towards CD3 (clone SK7, final dilution 1:25), CD19 (clone HIB19, 1:40), CD27 (clone L128, 1:200), CD38 (clone HB7, 1:20) and IgD (clone IA6-2, 1:33.3). T and B cells were identified among total lymphocytes by expression of CD3 and CD19, respectively. CD4 and CD8 T cells were quantified after staining of CD4 (clone SK3, 1:100) and CD8 (clone RPA-T8). Plasmablasts were identified by expression of CD38 among IgD-CD27+ CD19 positive switched-memory B cells. Analysis was performed on a BD FACSLyric flow-cytometer and BD FACSuite software v1.4.0.7047 followed by data analysis using FlowJo software 10.6.2. The gating strategy has been described before [9]. Absolute lymphocyte numbers were calculated based on differential blood counts.

### Preparation of UV-inactivated viral strains

Sample inactivation for further processing was performed with previously evaluated methods [36]. Briefly, SARS-CoV-2 stocks were aliquoted in small volumes (100 μl) and exposed to UV light in open vessels. The inactivation success was verified by TCID_50_ assay using A549-AT cells. The following SARS-CoV-2 isolates were used in this study: Parental strain (SARS-CoV-2 B.1 FFM7/2020, GenBank ID MT358643), Alpha (SARS-CoV-2 B.1.1.7 FFM-UK7931/2021, GenBank ID MZ427280.1), Beta (SARS-CoV-2 B.1.1.7 FFM-ZAF/2021, GenBank ID MW822592), Delta (SARS-CoV-2 B.1.617.2 FFM-IND8424/2021, GenBank ID MZ315141), BA.1 (SARS-CoV-2 B.1.1.529 FFM-SIM0550/2021 (EPI_ISL_6959871), GenBank ID OL800702), BA.2 (SARS-CoV-2

BA.2 FFM-BA.2-3833/2022, GenBank ID OM617939) [37-40]. For normalization, genome copy equivalents were determined using a one-step RT-digital PCR approach using the QIAcuity One-Step Viral RT-PCR Kit (Qiagen, Hilden, Germany) with a QIAcuity One Digital PCR System (Qiagen, Hilden, Germany). Primer and probe sequences (CDC N1/N2) were obtained from Integrated DNA Technologies (IDT).

### Quantification of vaccine-induced spike-specific T cells

SARS-CoV-2 specific T cells were determined from heparinized whole blood after stimulation exactly as described before [9, 15]. Stimuli included overlapping peptides spanning the SARS-CoV-2 spike protein (N-terminal receptor binding domain and C-terminal portion including the transmembrane domain, each peptide 2μg/ml; JPT, Berlin, Germany) with 0.64% DMSO and with 2.5μg/ml of *Staphylococcus aureus* enterotoxin B (SEB; Sigma) as negative and positive controls, respectively. In addition, we used titred amounts (22.5μl/225μl blood) UV-inactivated viral strains including medium as negative controls (Minimum Essential Medium (MEM) supplemented with 10% fetal calf serum (FCS), 4mM L-glutamine, 100IU/mL of penicillin, and 100μg/mL of streptomycin). All stimulations were carried out in presence of co-stimulatory antibodies against CD28 and CD49d (clone L293 and clone 9F10, 1μg/ml each). Immunostaining was performed using anti-CD4 (clone SK3, 1:33.3), anti-CD8 (clone SK1, 1:12.5), anti-CD69 (clone L78, 1:33.3), anti-IFNγ (clone 4S.B3, 1:100), anti-IL-2 (clone MQ1-17H12, 1:12.5), anti-TNFαα (clone MAb11, 1:20), and anti-CTLA-4 (clone BNI3, 1:50) and analyzed using flow-cytometry (BD FACS Canto II including BD FACSDiva software 6.1.3). SARS-CoV-2-reactive CD4 or CD8 T cells were identified as activated CD69-positive T cells producing IFNγ. Moreover, co-expression of IFNγ, IL-2 and TNFα was analyzed to characterize cytokine-expression profiles using a gating strategy as described before [9]. Reactive CD4 and CD8 T-cell levels after control stimulations were subtracted from levels obtained after SARS-CoV-2 specific stimulation, and 0.03% of reactive T cells was set as detection limit as described before [9].

### Determination of SARS-CoV-2 specific antibodies and neutralization capacity

Spike-specific IgG were determined using an enzyme-linked immunosorbent assay (ELISA, SARS-CoV-2-QuantiVac) as described before [9]. Antibody binding units (BAU/ml) <25.6 were scored negative, ≥25.6 and <35.2 were scored intermediate, and ≥35.2 were scored positive.

SARS-CoV-2 specific IgG towards the nucleocapsid (N) protein were quantified using the anti-SARS-CoV-2-NCP-ELISA. A surrogate neutralization assay (SARS-CoV-2-NeutraLISA) was used at a single serum dilution. Surrogate neutralizing capacity was calculated as percentage of inhibition (IH) by 1 minus the ratio of the extinction of the respective sample and the extinction of the blank value. A percentage of inhibition (IH) <20% was scored negative, IH≥20 and <35 intermediate, and IH≥35% positive. All assays were performed according to the manufacturer’s instruction (Euroimmun, Lübeck, Germany).

Antibody-mediated neutralization of authentic SARS-CoV-2 variants was performed as described before [40] using heat inactivated and serially diluted plasma samples (1:2). Samples were incubated with 4000 TCID50/mL of each SARS-CoV-2 variants. Infected cells were monitored microscopically for cytopathic effect formation 48 h post inoculation. Assays were performed testing each sample in a parallel approach comparing Delta and Omicron neutralization.

### Statistical analysis

Kruskal-Wallis test, followed by Dunn’s multiple comparisons test, was performed to compare unpaired non-parametric data such as lymphocyte subpopulations, T-cell and antibody levels, and CTLA-4 expression. Data with normal distribution such as cytokine-expression profiles, age were analyzed using ordinary one-way ANOVA. Categorial analyses on gender, vaccine responses, and adverse events were performed using X^2^ test. Correlations between levels of T cells towards overlapping peptides and UV-inactivated viral strains were analyzed by correlation matrix according to Spearman. A p-value <0.05 was considered statistically significant. Analysis was carried out using GraphPad Prism 9.4.1 software (GraphPad, San Diego, CA, USA) using two-tailed tests.

## Data Availability

All figures and tables have associated raw data. The data that support the findings of this study are available from the corresponding author upon request.

## Acknowledgements

The authors thank Dr. Christina Baum from the occupational health care center at Saarland University Medical Center, and Dr. Wilhelm Walch and Dr. Dirk Jesinghaus from the vaccine center in Neunkirchen for their support in enrolling participants. Furthermore, we would like to thank Christiane Pallas for excellent technical assistance. The authors also thank all participants to this study. Financial support was provided by the State chancellery of the Saarland to M.S. Moreover, the work was in part supported by the Clusterproject ENABLE and The Innovation Center TheraNova funded by the Hessian Ministry for Science and the Arts and the German Federal Ministry of Education and Research (BMBF) funding to M.W. (COVIDready, grant number 02WRS1621C).

## Author Contributions

T.S., U.S., and M.S. designed the study; F.H., V.K., T.S., U.S., M.W. and M.S. designed the experiments, F.H., V.K., S.M., A.A.-O., L.Z., C.G., R.U., A.W., M.W. and T.S. performed experiments; F.H. and U.S., and M.S. contributed to study design, patient recruitment, and clinical data acquisition. F.H. T.S., U.S. and M.S. performed statistical analysis. T.S., U.S., and M.S. supervised all parts of the study; F.H., T.S., M.W. and M.S. wrote the manuscript. All authors approved the final version of the manuscript.

## Supplementary figure

**Supplementary figure S1:**
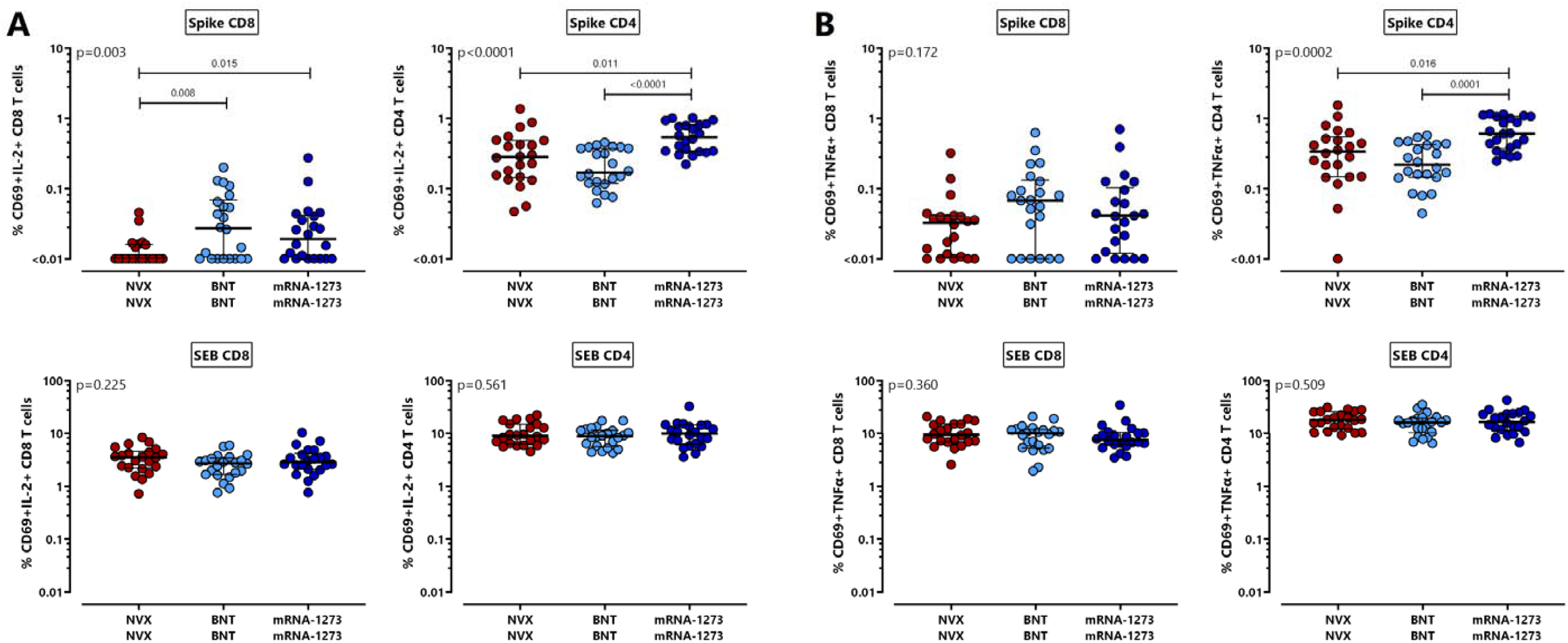
Levels of spike-specific T cells producing IL-2or TNFαα. Spike-specific CD8 and CD4 T cells were quantified and characterized by intracellular cytokine staining after antigen-specific stimulation of whole blood samples of 66 individuals 13-18 days after the second vaccination with NVX-CoV2373 (NVX), BNT162b2 (BNT) or mRNA-1273 (n=22 each). Spike-specific and SEB-reactive CD8and CD4T cells were determined based on co-expression of CD69 and **(A)** IL-2 or **(B)** TNFαα with respective reactivity after control stimulation subtracted. Bars represent medians with interquartile ranges. Differences between the groups were calculated using two-sided Kruskal-Wallis test with Dunn’s multiple comparisons post-test. IL, interleukin; SEB, Staphylococcus aureus enterotoxin B; TNFα, tumor necrosis factor.

